# Field-deployable, rapid diagnostic testing of saliva samples for SARS-CoV-2

**DOI:** 10.1101/2020.06.13.20129841

**Authors:** Shan Wei, Esther Kohl, Alexandre Djandji, Stephanie Morgan, Susan Whittier, Mahesh Mansukhani, Raymond Yeh, Juan Carlos Alejaldre, Elaine Fleck, D’Alton Mary, Yousin Suh, Zev Williams

## Abstract

Rapid, scalable, point-of-need, COVID-19 diagnostic testing is necessary to safely re-open economies and prevent future outbreaks. We developed an assay that detects single copies of SARS-CoV-2 virus directly from saliva and swab samples in 30 min using a simple, one-step protocol that utilizes only a heat block and microcentrifuge tube prefilled with a mixture containing the necessary reagents and has a sensitivity and specificity of 97% and 100%, respectively.

---

To safely re-open economies closed due to the COVID-19 pandemic as well as to prevent future outbreaks, diagnostic testing capacity must be massively expanded ^1, 2^. Additionally, diagnostic tests must be rapid and available in the field, such as prior to boarding a flight or entering a nursing home^2^. Nucleic acid tests (NAT) that utilize RT-PCR, isothermal amplification, and Crispr-Cas12 are the standard methods for detection of SARS-CoV-2 from swab and saliva specimens ^3-5^. However, due to inhibitors present in both transport media and, even more so, saliva, these NAT-based methods require RNA extraction or sample pretreatment prior to amplification. Consequently, samples must be either transported to centralized high-complexity laboratories or processed at the point-of-care using systems that rely on specialized, proprietary instruments and consumables, increasing costs and limiting the capacity to scale and implement widespread testing both in the U.S. and globally ^6, 7^.

Here we report the development and initial validation of a one-step, SARS-CoV-2 detection assay that can detect single-copy levels of virus directly from saliva using only a 1.5 mL microcentrifuge tube preloaded with a reaction mixture and a 30 min incubation in a heat block, without the need for RNA extraction or sample pretreatment (Figure 1a). The method is based on significant modifications to <u>R</u>everse <u>T</u>ranscription <u>L</u>oop-mediated isothermal <u>Amp</u>lification (RT-LAMP), a targeted nucleic acid amplification method that utilizes a combination of 2-3 primer sets and a DNA polymerase with high strand displacement activity ^8^. Because RT-LAMP is severely inhibited by saliva, significant modifications were necessary to increase the sensitivity of the assay in saliva by >1000-fold. We term the new assay <u>H</u>igh-<u>P</u>erformance LAMP (HP-LAMP).

**Figure 1.**
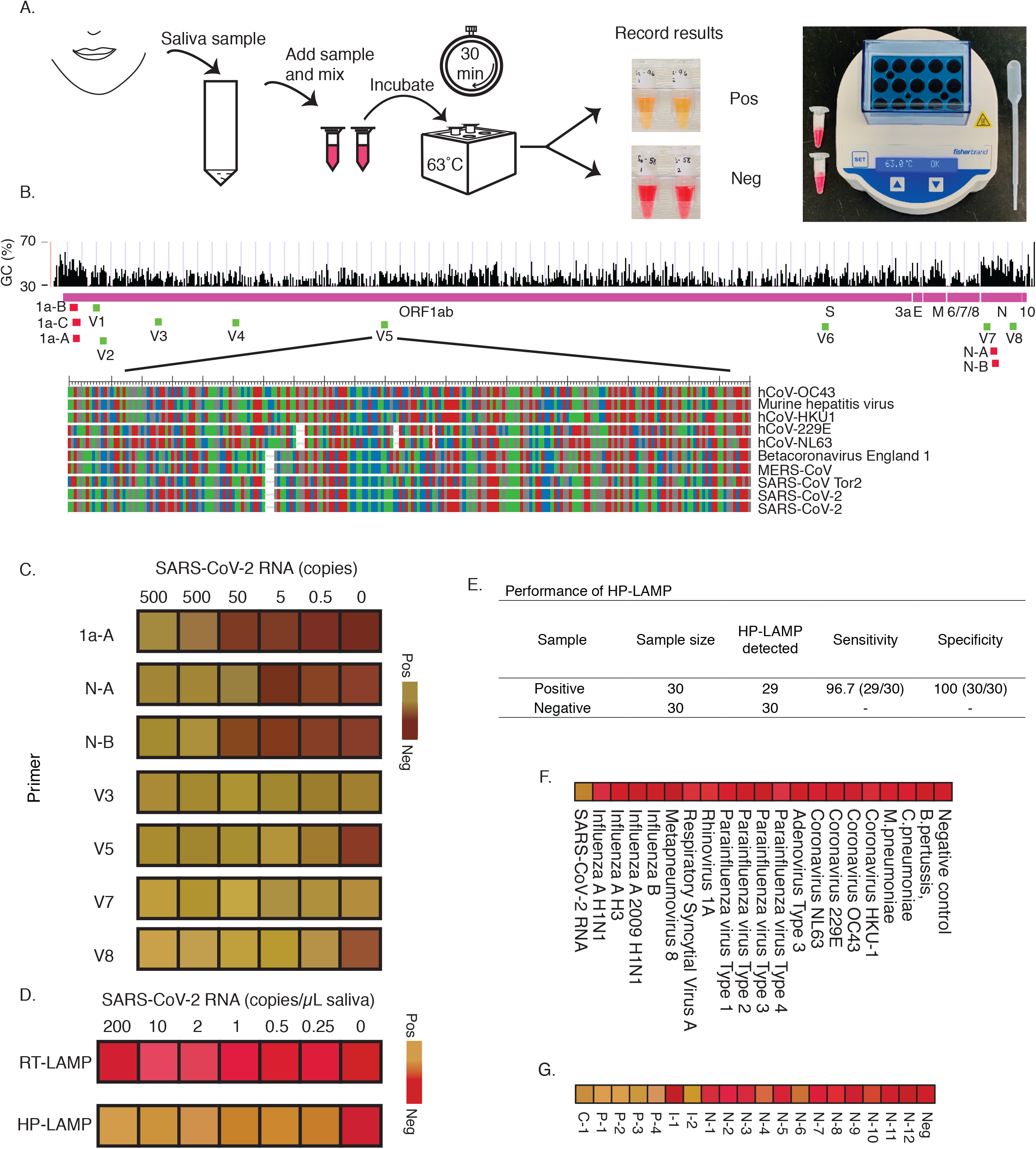
**A)** Schematic of workflow for SARS-CoV-2 testing directly from saliva using HP-LAMP. Saliva may be directly collected from the mouth or first collected in a container. Saliva was added to 1.5 ml microcentrifuge tubes pre-filled with the reaction mixture, incubated for 30 min at 63 □C and then visualized for colorimetric change (yellow=positive; red=negative). The minimum equipment needed to run the assay is a disposable transfer pipette, heat block, and microcentrifuge tubes prefilled with reaction mixture. No prior RNA extraction or treatment is necessary. Testing performed in the appropriate safety environment. **B)** Genome map showing targeted region of primers. Primers from previous publication are indicated in red (1a-A, 1a-B, 1a-C, N-A, N-B). In-house designed primer sets are indicated in green (V1-8). The targeted region of primer set V5 with alignments of other Betacoronavirus genomes are featured. Each nucleic acid is shown (A: green; G: gray; T: red; C: blue). The percentage of GC-content across the genome is indicated. **C)** Screening primer sets for SARS-CoV-2 testing. Those primer sets with sensitivities of 500 copies or fewer of SARS-CoV-2 are shown. A negative control and between 0.5 – 500 copies of SARS-CoV-2 RNA were spiked into 25 μL reaction volume and assayed using RT-LAMP with previously reported primer sets (1a-A, N-A, N-B) and in-house designed primer sets (V3, V5, V7, V8). Color blocks reflect the actual color captured from a 96-well PCR plate. **D)** Comparison of RT-LAMP and HP-LAMP sensitivity. Contrived saliva samples containing 0.25-200 copies of SARS-CoV-2 viral RNA per μL of saliva, along with a negative control, were tested using the V5 primer set with RT-LAMP and HP-LAMP. While RT-LAMP failed to detect viral RNA at levels as high as 200 copies/ μL of saliva, HP-LAMP consistently detected levels as low as 1 copy/μL of saliva and often less. **E)** Performance characteristics of HP-LAMP. A total of 60 contrived samples (30 SARS-CoV-2 positive and 30 SARS-CoV-2 negative) were evaluated using HP-LAMP. Positive samples contained 2 copies of SARS-CoV-2 viral RNA per μL of saliva. **F)** HP-LAMP testing on respiratory verification panel of 19 respiratory virus and bacteria and SARS-CoV-2 viral RNA. **G)** HP-LAMP testing of saliva prospectively collected from patients undergoing nasopharyngeal swab sampling. Of the 149 nasopharyngeal samples collected, 4 tested positive, 2 tested indeterminate and the remainder tested negative. The corresponding saliva samples were tested using HP-LAMP. Saliva from all 4 patients with SARS-CoV-2 positive nasopharyngeal swab samples (P-1, P-2, P-3, P-4) tested positive with HP-LAMP. Saliva from 1/2 patients with indeterminate SARS-CoV-2 nasopharyngeal swab samples (I-2) tested positive with HP-LAMP and 1/2 (I-1) tested negative. Saliva from all 12 patients with negative nasopharyngeal swab samples (N-1 to N-12) and the negative control (Neg) tested negative with HP-LAMP. The positive control (C-1) was a contrived sampled consisting of 5 copies of SARS-CoV-2 viral RNA per μL of saliva. For panels **C, D, F** and **G**, color blocks reflect the actual color of assay tubes. (yellow=positive; red=negative)

We first designed eight sets of six primers targeting regions across the full length of the SARS-CoV-2 genome (Figure 1B). Typically, primers for LAMP are designed to target GC-rich regions of the viral RNA because GC-rich regions bind more tightly to primers. However, in SARS-CoV-2, these regions are found towards the 5′ and 3′ ends of the viral RNA. Because salivary nucleases degrade viral RNA from the 3′ and 5′ ends, we reasoned that the central portion of the virus would be better protected and, therefore, designed our primers to target that region. In the case of SARS-CoV-2, the central region is GC-poor (AT-rich), making it difficult to select primer candidates with optimal annealing temperatures when following standard parameters for primer design. Therefore, we designed the primers to permit large primer-mediated loop-structures while ensuring that the primers did not form stable secondary structures or self-dimerize. We also aligned the known SARS-CoV-2 genomic sequence with those of six other human coronaviruses (SARS-CoV, MERS-CoV, HCoV-HKU-1, HCoV-NL63, HCoV-OC43 and HCoV-229E) to ensure no cross-reactivity. To determine which primer set was most sensitive and specific to SARS-CoV-2, we tested the eight primer sets that we designed, along with previously published primer sets ^9, 10^, using serial dilutions of 500 to 0.5 copies of SARS-CoV-2 RNA standard spiked into a 25 μL RT-LAMP reaction (Figure 1C). Primer set V5 detected 10^0^ to 10^−1^ level viral RNA in water, representing a 10- to 100-fold improvement in sensitivity and equivalent specificity compared with previously published primer sets. Additionally, primer set V5 targeted 3640 out of 3672 (>99%) complete SARS-CoV-2 genomes archived on the NCBI Virus database with no mismatches (as of June 9, 2020), and was, therefore, selected for use in our assay.

Even with the improved primer sets, the RT-LAMP reaction was still not sufficiently sensitive to detect fewer than 200 viral copies/μL in saliva, which is far higher than the ≤ 2 viral copies/μL limit considered necessary for testing clinical samples (Figure 1D). In order to achieve the necessary > 100-fold improvement in sensitivity while maintaining a 100% specificity, we systematically modified the RT-LAMP reaction conditions to improve performance. We found that sensitivity and specificity of the assay could be markedly improved by adding carrier DNA, carrier RNA, and RNase inhibitors, as well as by increasing the reaction volume (Supple. Figure 1A-1E). Because our assay was so sensitive, there was a risk that carry-over product from prior samples could cross contaminate a new sample and lead to false-positive results. To solve this problem, we added dUTP to our reaction mixture in order for it to be incorporated into the HP-LAMP product. We also added Antarctic Thermolabile uracil-DNA N-glycosylase (UDG), which degrades any dUTP-containing product from prior reactions but is itself inactivated at temperatures below our running conditions, to the final HP-LAMP reaction mixture (Supple. Method).

To determine the analytical limit of detection (LoD) of HP-LAMP, 0.25-200 copies of SARS-CoV-2 RNA per μL of saliva were tested using both RT-LAMP and HP-LAMP (Figure 1D). While RT-LAMP was unable to detect 200 or fewer copies of viral RNA per μL of saliva, HP-LAMP was able to consistently detect 1 copy of viral RNA per μL of saliva (10/10). We, therefore, set the clinical LoD to double this amount (2-fold LoD), or 2 copies of viral RNA per μL of saliva. Since viral transport medium (VTM) is less inhibitory to RT-LAMP than saliva, HP-LAMP was able to detect 10^0^ to 10^−1^ level viral RNA spike-in in VTM, making it a promising versatile detection method for saliva and swab samples (Supple. Figure 1F).

We then assessed the sensitivity and specificity of our reaction. We tested thirty contrived positive samples consisting of saliva from COVID-19 negative individuals with 2 copies of SARS-CoV-2 per μL of saliva spiked-in, and thirty negative samples consisting of saliva samples from COVID-19 negative individuals without viral RNA spiked-in. Of the positive samples, 29/30 were detected and 30/30 of the negative samples remained negative for a sensitivity of 97% and a specificity of 100% (Figure 1E).

To determine if there was cross reactivity of our assay with other known respiratory viruses, we tested 19 known respiratory viruses and bacteria spiked-in to saliva from healthy individuals as well as a positive control consisting of 5 copies of SARS-CoV-2 spike-in per μL of saliva. No cross reactivity with other respiratory pathogens was noted, while a positive signal was obtained from the SARS-CoV-2 spike-in sample (Figure 1F).

We next determined the ability of our assay to accurately detect SARS-CoV-2 directly from patient saliva samples. This portion of our study was reviewed and approved by the Columbia University IRB (#AAAS9893) and all methods were carried out in accordance with relevant guidelines and regulations. All study subjects signed informed consent prior to participating. From 04/29/2020 to 06/1/2020 we prospectively collected saliva samples at the time when patients presented to Columbia University Irving Medical Center’s COVID-19 testing tent and cough/fever clinic for clinical SARS-CoV-2 testing via nasopharyngeal swab. Nasopharyngeal swab samples were tested on the RT-PCR-based Roche Cobas 6800 system following routine laboratory protocols according to the manufacturer’s recommendations. Paired saliva samples were tested using the HP-LAMP assay. During that time, a total of 149 nasopharyngeal and paired saliva samples were collected and archived for use in this validation. All nasopharyngeal swab samples were tested using the Roche RT-PCR System. Of these, only 4 tested positive, consistent with the decreasing rate of new cases in New York City during that time and the 4% positivity rate seen city-wide ^11^. Of the remaining samples, 2 tested indeterminate, and 143 tested negative. We then tested saliva from the 4 positive, 2 indeterminate and 12 randomly selected negative samples using HP-LAMP as well as undergoing RNA extraction and RT-PCR. Results using HP-LAMP were concordant with the nasopharyngeal swab results for all 4 positive and 12 negative samples. Of the indeterminate results, one tested positive and one tested negative with HP-LAMP (Figure 1G, Supple. Table). In addition, we extracted RNA from the selected saliva samples and tested using RT-PCR following New York SARS-CoV-2 Real-time Reverse Transcriptase (RT)-PCR Diagnostic instructions. The Ct values for the 4 positive samples ranged from 25.3-34 (Target N1), 26.3-36 (Target N2), corresponding to 2.2-1334 copies/µL saliva. No virus was detected in either of the indeterminate nor in the negative samples (Supple Table).

In summary, we developed HP-LAMP, which enables rapid detection of SARS-CoV-2 directly from saliva in 30 min using a simple one-step protocol with a LoD of 2 viral copies per μL of saliva and a sensitivity and specificity of 97% and 100%, respectively. Performing the assay requires only a heat block and a 1.5 mL microcentrifuge tube prefilled with a mixture containing the necessary enzymes, primers, buffers and reagents to simultaneously perform reverse transcription and amplification of the SARS-CoV-2 viral RNA, while blocking the naturally occurring inhibitors and nucleases found in saliva. This simple method can be easily scaled and deployed to centralized laboratories, points-of-care and field locations where testing is greatly needed.

## Data Availability

No sequencing data was generated in this study.

## Acknowledgement

We thank Drs. Kevin Roth, Steven Spitalnik, and Eldad Hod of the Department of Pathology and Cell Biology and members of the Williams Laboratory, Columbia University Fertility Center, Suh Laboratory, Clinical Microbiology Laboratory, Columbia University Laboratory of Personalized Genomic Medicine at Columbia University Medical Center and New York Presbyterian Hospital for their helpful inputs and support for this study. This study is supported by NIH grants R01HD100013 (Z.W.), R01HD086327 (Z.W.), 1RF1AG057341 (Y.S), and R01AG057433 (Y.S).

---

**Table 1.** Comparison of detection methods of SARS-CoV-2

|  | **Xpert**<br>**Xpress**<br>**SARS-CoV-**<br>**2 test** | **Roche**<br>**Cobas** | **SARS-CoV-2**<br>**DETECTR**<br>**assay** | **HP-LAMP for**<br>**SARS-CoV-2** |
| --- | --- | --- | --- | --- |
| Technique | RT-PCR | RT-PCR | RT-LAMP +<br>CRISPR–<br>Cas12 | Colorimetric<br>RT-LAMP |
| Target | E gene, N<br>gene | E gene, N<br>gene | E gene, N<br>gene | Orf1ab |
| Sample type | Swabs | Swabs | Swabs | Saliva |
| RNA extraction | Yes | Yes | Yes | No |
| Time for assay | 45min | 3.5h | 45* - 100min | 30min |
| Detection<br>sensitivity<br>(copies/μL) | 0.250 | 0.10-0.15 | 10 | 1-2 |
| Bulk equipment | Yes | Yes | No | No |

